# Brain growth charts of “clinical controls” for quantitative analysis of clinically acquired brain MRI

**DOI:** 10.1101/2023.01.13.23284533

**Authors:** Jenna M. Schabdach, J. Eric Schmitt, Susan Sotardi, Arastoo Vossough, Savvas Andronikou, Timothy P. Roberts, Hao Huang, Viveknarayanan Padmanabhan, Alfredo Oritz-Rosa, Margaret Gardner, Sydney Covitz, Saashi A. Bedford, Ayan Mandal, Barbara H. Chaiyachati, Simon R. White, Ed Bullmore, Richard A.I. Bethlehem, Russell T. Shinohara, Benjamin Billot, J. Eugenio Iglesias, Satrajit Ghosh, Raquel E. Gur, Theodore D. Satterthwaite, David Roalf, Jakob Seidlitz, Aaron Alexander-Bloch

## Abstract

**Background:** Brain MRIs acquired in clinical settings represent a valuable and underutilized scientific resource for investigating neurodevelopment. Utilization of these clinical scans has been limited because of their clinical acquisition and technical heterogeneity. These barriers have curtailed the interpretability and scientific value of retrospective studies of clinically acquired brain MRIs, compared to studies of prospectively acquired research quality brain MRIs.

**Purpose:** To develop a scalable and rigorous approach to generate clinical brain growth chart models, to benchmark neuroanatomical differences in clinical MRIs, and to validate clinically-derived brain growth charts against those derived from large-scale research studies.

**Materials and Methods:** We curated a set of clinical MRI *S*cans with *L*imited *I*maging *P*athology (SLIP) – so-called “clinical controls” – from an urban pediatric healthcare system acquired between 2005 and 2020. The curation process included manual review of signed radiology reports, as well as automated and manual quality review of images without gross pathology. We measured global and regional volumetric imaging phenotypes in the SLIP sample using two alternative, advanced image processing pipelines, and quantitatively compared clinical brain growth charts to research brain growth charts derived from >123,000 MRIs.

**Results:** The curated SLIP dataset included 372 patients scanned between the ages of 28 days post-birth and 22.2 years across nine 3T MRI scanners. Clinical brain growth charts were highly similar to growth charts derived from large-scale research datasets, in terms of the normative developmental trajectories predicted by the models. The clinical indication of the scans did not significantly bias the output of clinical brain charts. Tens of thousands of additional healthcare system scans meet inclusion criteria to be included in future brain growth charts.

**Conclusion:** Brain charts derived from clinical-controls are highly similar to brain charts from research-controls, suggesting that curated clinical scans could be used to supplement research datasets.

**Summary Statement:** Brain growth charts of pediatric clinical MRIs with limited imaging pathology (N=372) are highly correlated with charts from a large aggregated set of research controls (N>120,000).

**Key Results:** A cohort of brain MRI scans with limited reported imaging pathology (N=372, 186 female; ages 0.07 - 22.2 years, median = 10.2) were identified using signed radiology reports and processed using two segmentation pipelines. Growth charts generated from these scans are highly correlated with growth charts from a large aggregated set of research controls (r range 0.990 - 0.999). There was no evidence of bias due to the reason for each scan.

## Introduction

A principal challenge in brain MRI research is recruiting large numbers of participants necessary to support valid scientific inference and generalizability. Retrospective studies of clinically acquired brain MRIs may help supplement costly prospective neuroimaging studies by harnessing existing healthcare systems data. For example, individuals with specific diseases may lack the support or resources to participate in research studies, but some of their clinical data could be anonymized for secondary use in research applications. From this perspective, the millions of brain MRIs acquired each year in clinical settings represent a valuable and vastly underutilized resource.

However, a major obstacle for studies using clinically-acquired MRI is the lack of appropriate “control groups,” which are necessary to rigorously test hypotheses in a patient group of interest. In research settings, it is customary to recruit “healthy”, “cognitively normal”, or “unaffected” participants explicitly for this purpose. For clinically-acquired MRIs, a control group might be composed of demographically-matched patients who underwent brain MRIs to rule out serious neuropathology and were found to have unremarkable MRIs (Kerr et al 2022). A critical and unresolved question is whether the difference in ascertainment process between research-controls and clinical-controls biases inferences about patient groups of interest.

Other challenges in using clinically-acquired brain MRI data include the differences due to the use of multiple scanners and the variability in the quality of scans. Recent statistical approaches have proven successful in harmonizing scanner and sequence differences in MRI data (Johnson and Li 2007; Fortin et al 2017; Pomponio et al 2019). Additionally, deep-learning based tools may provide robustness to scan quality during image segmentation (Billot et al. 2022) and assessment of scan quality (McClure et al 2019). Recently, we developed research-control brain growth charts to quantitatively benchmark brain MRI phenotypes against population norms while also controlling for technical differences between sites in a large aggregated neuroimaging dataset (Bethlehem, Seidlitz, and White et al 2022).

In the present study, we sought to evaluate if a cohort of clinically-imaged controls had demonstrable brain development differences from research-controls. First, we identified a large set of scans with limited imaging pathology (SLIP) with clinically acquired brain MRIs provided by an urban pediatric healthcare system. We then quantitatively compared clinical brain growth charts to research brain growth charts derived from >123,000 MRIs included in the Lifespan Brain Chart Consortium (LBCC; Bethlehem, Seidlitz, and White et al. 2022). We evaluate the similarities between the clinical-controls and the research-controls in terms of growth trajectories and age at peak cortical region volumes.

## Materials and Methods

This study was reviewed by the IRB and determined to be exempt from further oversight because it consisted of secondary analysis of preexisting clinical data. The remainder of this section contains an abbreviated overview of our methods. Please see the Supplementary Materials for an extended description.

### Dataset curation

Aggregating and curating the SLIP dataset began with a request for 1013 scan sessions determined by manual review of their signed radiology reports to lack significant clinical pathology. These scan sessions were requested from the radiology department with the aid of an honest broker. This request was limited to 3 Tesla (3T) scans, as the department deployed a harmonized MPRAGE T1-weighted (T1w) sequence for routine brain MRIs across its 3T clinical scanners beginning in 2008 (see Supplementary Materials). The scans were organized into BIDS format (Gorgolowski et al 2016) using *heudiconv* (Halchenko et al 2020) and filtered according to their metadata using *CuBIDS* (Covitz et al 2022) to isolate non-contrast high-resolution T1w scans (<=1×1×1mm; N=444 scans), then were manually graded by two raters to remove low-quality scans (e.g, scans with significant motion artifact) (Rosen et al 2018; Bedford et al 2022) (N=372 scans). An overview of data curation and quality control is provided in Figure 1 with complete details in Supplemental Materials.

**Figure 1.**
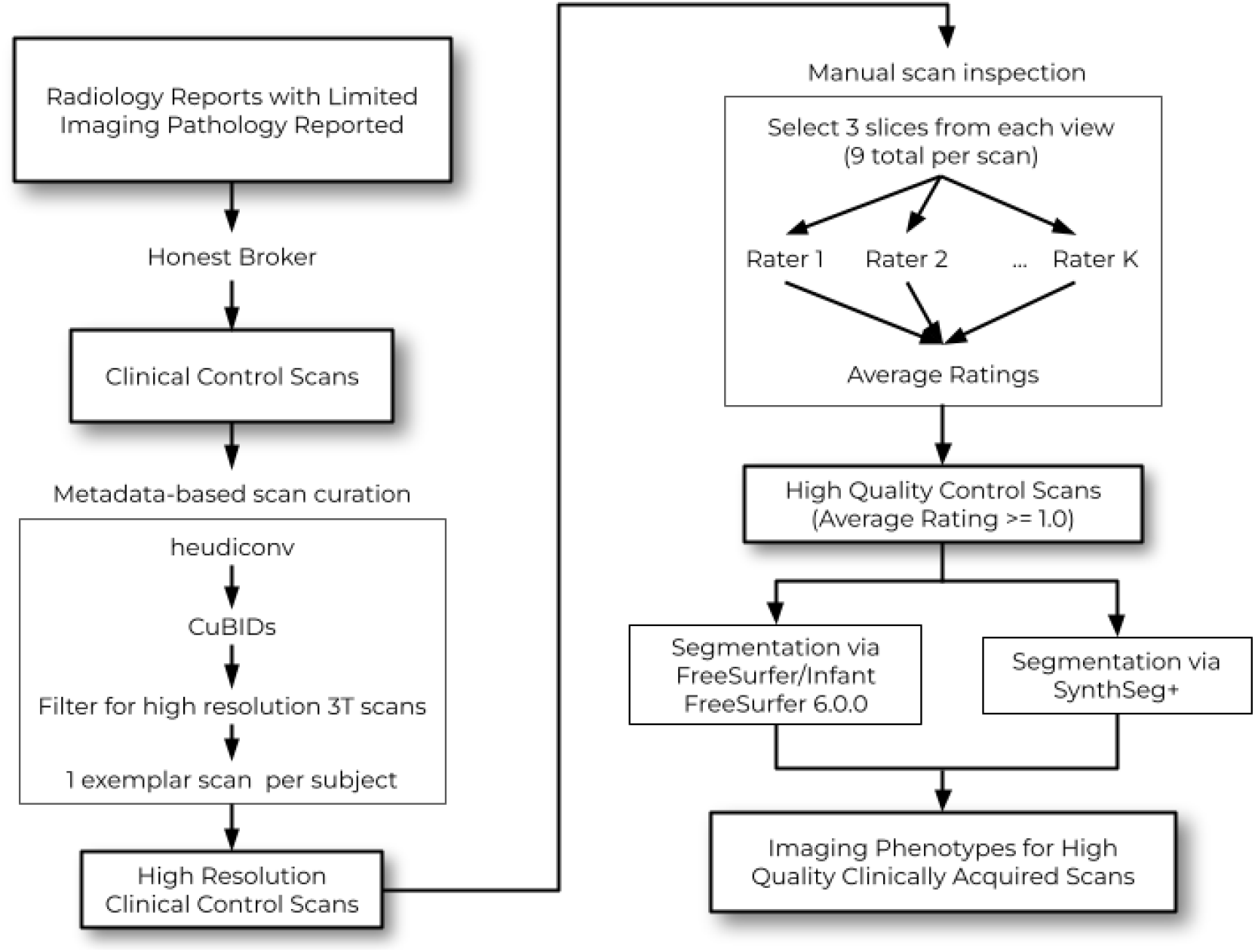
Overview of the data curation and processing pipeline. The initial request for sessions whose signed radiology reports contained no gross pathology was submitted to the honest broker. The honest broker returned a set of anonymized MRI scans which were then filtered to identify only high resolution T1 weighted scans from 3T scanners. Next, the high resolution scans were manually reviewed by independent raters to remove visually low quality images. This finalized set of SLIP scans was processed using two neuroimaging processing pipelines (FreeSurfer/Infant FreeSurfer 6.0.0 and SynthSeg+) to produce two sets of imaging phenotypes for high-quality clinically acquired scans.

### Image Processing

Two parallel processing pipelines were used for quantitative analyses of T1w scans. The first pipeline reoriented and aligned scans to the MNI152 atlas (Fonov et al 2011), removed facial features, and performed segmentation using either FreeSurfer 6.0.0 (for patients > 3 years old) (Jenkinson et al 2012; Fischl et al 2004) or Infant FreeSurfer 6.0.0 (for patients <= 3 years old) (Zöllei et al 2020). The second pipeline performed segmentation using SynthSeg+ with pretrained models (Billot et al 2022). Both pipelines produced quantitative volumetric measurements subsequently referred to as “imaging phenotypes”. Global imaging phenotypes quantified by each pipeline included total cortical gray matter volume (GMV), white matter volume (WMV), subcortical gray matter volume (sGMV), ventricular volume (CSF), and total cerebrum volume (TCV). Additionally, 34 regional cortical volumes were quantified by the FreeSurfer pipeline using the sulcal-based Desikan-Killiany parcellation (Desikan et al 2006).

### Statistical Analysis

We sought to compare clinical brain growth charts to a subset of the previously published LBCC reference dataset of 123,984 MRIs aggregated across 100 primary studies (www.brainchart.io) (Bethlehem, Seidlitz, and White et al. 2022). The subset was limited to LBCC subjects processed using either FreeSurfer 6.0.0 or Infant FreeSurfer in the same age range as SLIP. Prior to growth chart modeling, ComBat, a batch correction approach adapted from statistical genomics, was used to harmonize the SLIP imaging phenotypes across scanners (Johnson and Li 2007; Fortin et al 2017; Pomponio et al 2019). ComBat was used to control for the effect of MR scanners while preserving the effect of other model covariates.

Growth charts of both SLIP and age-limited LBCC data were fit using generalized additive models for location, scale, and shape (GAMLSS): a distributional regression approach that models the mean, variance, and higher-order statistical moments in terms of flexible nonlinear associations using covariates of interest (Stasinopoulos et al 2017). A generalized gamma distribution was used to link each imaging phenotype to predictor variables using the GAMLSS package in R. The predictor variables included age and sex as reported in the electronic health record. For the FreeSurfer pipeline, the Euler number, a robust marker of image quality, was also included (Rosen et al. 2018). As detailed previously for LBCC growth charts, the mean and variance of each GAMLSS was modeled by a non-linear age effect using third-order fractional polynomials while an intercept term alone was employed for model skewness allowing for shared skewness across the age range.

## Results

### Demographics and quality control

The curated SLIP dataset consisted of 372 subjects (185 females) scanned between the ages of 28 days post-birth and 22.2 years during the time period of 2005 to 2020 (Table 1) across nine 3T MRI scanners (see Supplementary Table 2 for scanner details). There was no evidence of age or sex biases relating to specific scanners or to manual or automated measures of image quality (Figure 2). There were no statistical biases related to the year of the scan (Supplemental Figure 1). There was evidence of subtle but significant scanner effects on imaging phenotypes, which were mitigated using ComBat harmonization (Figure 3). The age-limited LBCC data used for global imaging phenotype comparison was taken from 23 studies with 8,346 subjects (3,399 females) between the ages of 152 days post-birth and 22 years (Supplemental Table 2). The publicly available LBCC growth charts were used as the baseline of comparison for the cortical region phenotypes.

**Table 1.** Demographic characteristics of curated SLIP dataset. The distribution of the year of scan is an artifact of the order in which radiology reports have been examined thus far.

|  | Total | Male | Female |
| --- | --- | --- | --- |
| Total | 372 | 187 | 185 |
| Age (years) |  |  |  |
| 0-2 | 41 | 25 | 16 |
| 2-5 | 41 | 22 | 19 |
| 5-10 | 102 | 51 | 51 |
| 10-13 | 63 | 37 | 26 |
| 13-18 | 113 | 44 | 69 |
| 18+ | 12 | 8 | 4 |
| Primary Reason for Scan |  |  |  |
| Developmental disorder | 22 | 13 | 9 |
| Clinical eye or vision finding | 31 | 18 | 13 |
| Headache | 156 | 67 | 89 |
| Suspected seizure | 33 | 18 | 15 |
| Other | 130 | 71 | 59 |
| Year of Scan |  |  |  |
| 2005 | 24 | 13 | 11 |
| 2006 | 58 | 26 | 32 |
| 2007 | 63 | 34 | 29 |
| 2008 | 66 | 34 | 32 |
| 2009 | 61 | 33 | 28 |
| 2010 | 78 | 38 | 40 |
| 2011+ | 22 | 9 | 13 |
| Race (as reported in EMR) |  |  |  |
| American Indian or Alaska Native | 1 | 1 | 0 |
| Asian | 7 | 4 | 3 |
| Black or African American | 63 | 33 | 30 |
| Multiple Races | 2 | 1 | 1 |
| Other | 32 | 19 | 13 |
| White | 267 | 129 | 138 |

**Figure 2.**
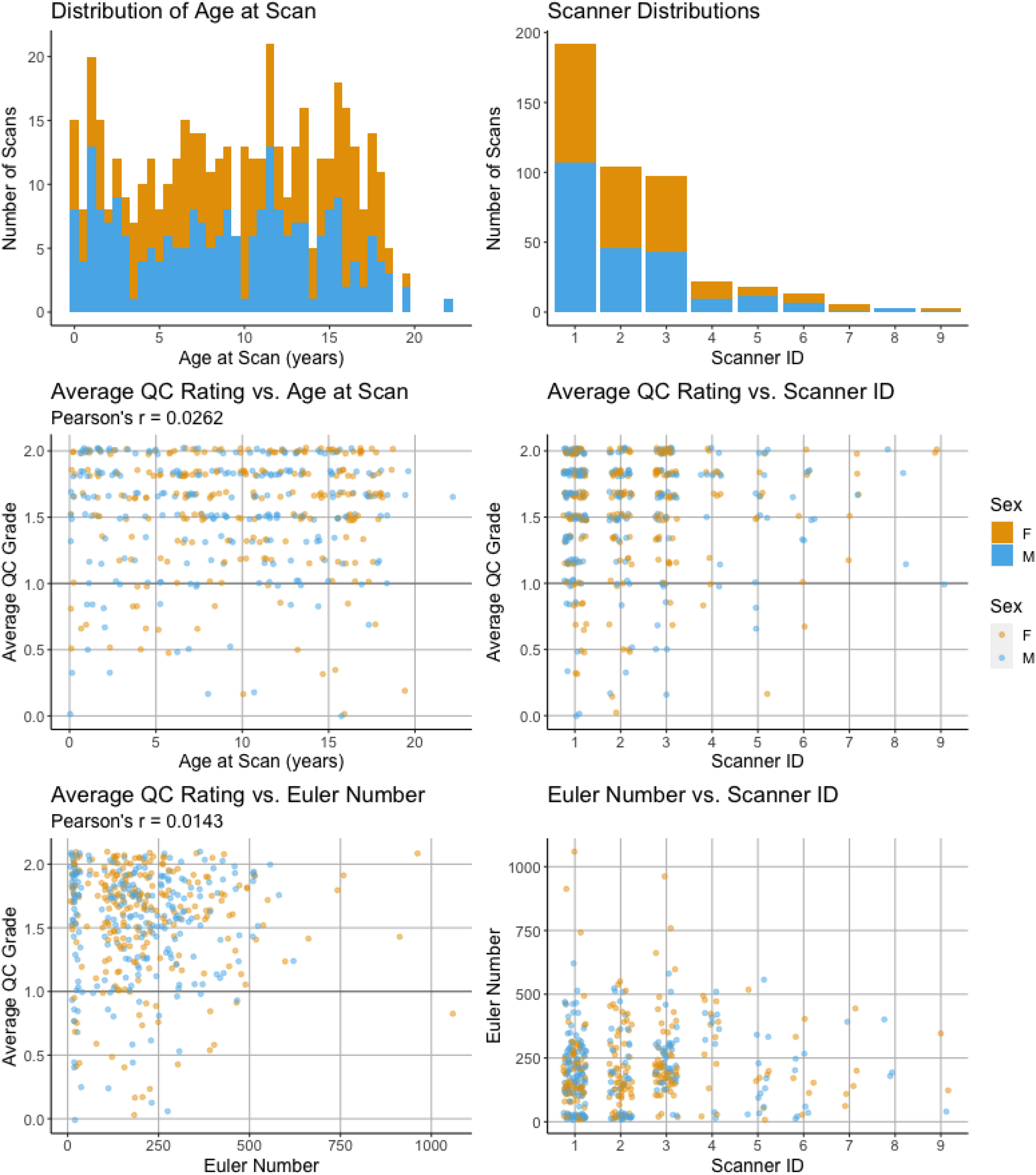
The distributions of patient ages, MRI scanner, and image quality in the curated SLIP dataset. The average quality control (QC) rating refers to an ordinal scale applied by two manual raters (0, poor quality; 1, acceptable quality; 2, highest quality). The Euler number refers to the number of holes in the FreeSurfer cortical surface reconstruction of the scan prior to topological correction. Scanner details are provided in Supplemental Table 1.

**Figure 3.**
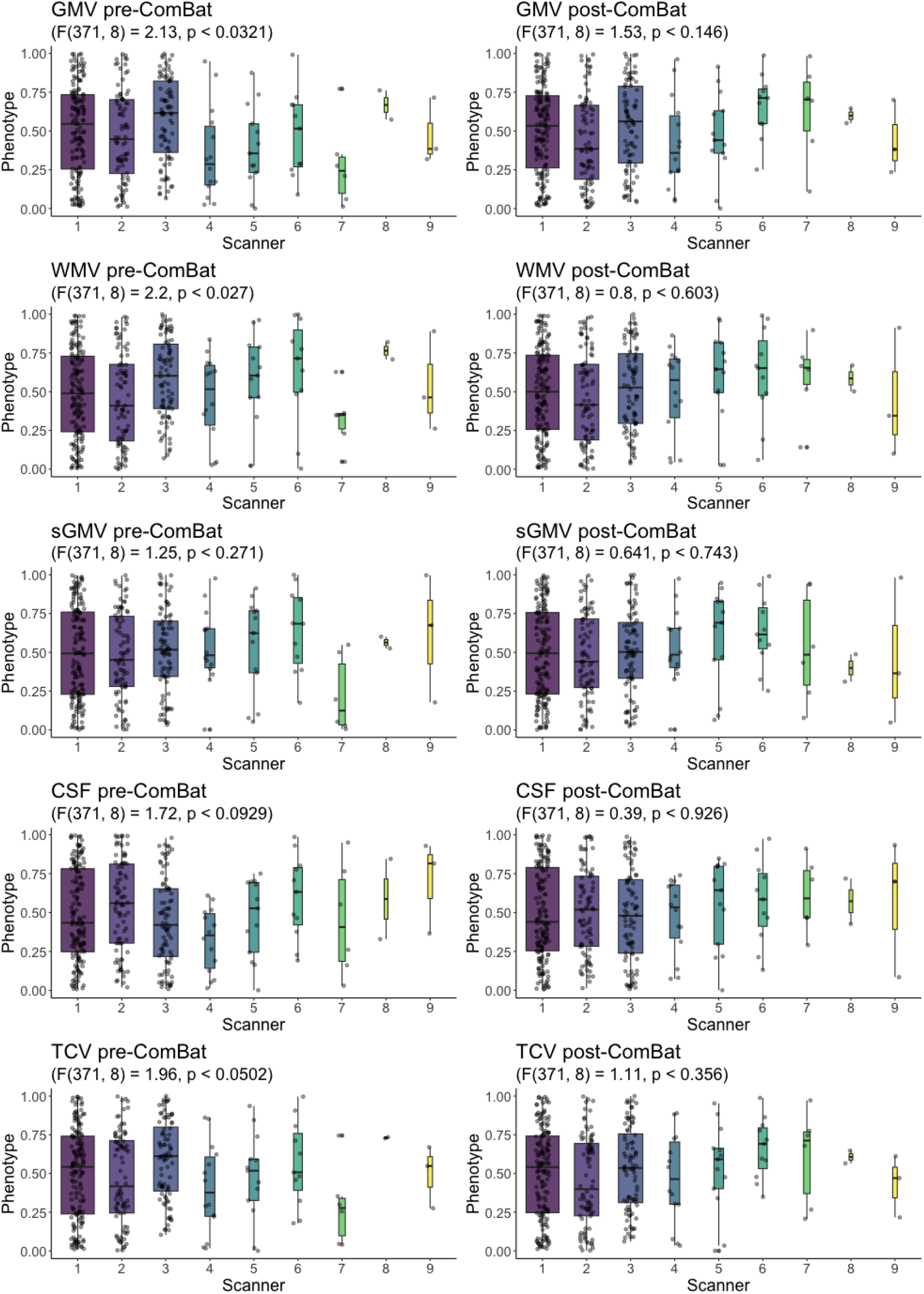
Centiles of global phenotypes across MRI scanners, before and after harmonization with ComBat across scanner ID. Boxplots of centiles from each scanner are represented individually and the scanners are presented left to right in decreasing order of the number of scans. The left column shows the centiles of FreeSurer (FS) imaging phenotypes prior to ComBat harmonization (pre-ComBat), while the right column shows the centiles of FS after ComBat (post-ComBat). The effect of the scanner was tested with ANOVA, showing no significant difference after harmonization. GMV, gray matter volume; WMV, white matter volume; sGMV, subcortical gray matter volume; CSF, ventricle volume; TCV, total cerebrum volume.

### Hospital growth charts have similar properties to research growth charts

We compared the characteristics of the SLIP and age-limited LBCC brain growth chart models to assess the validity of SLIP charts and the impact of the image processing pipeline (Figure 4). The charts display similar key milestones and overall developmental trajectory shapes, demonstrating robustness to the dataset and the choice of image processing pipeline. In general, the brain growth charts were highly similar between LBCC and SLIP (median phenotype trajectory correlation, r=0.997; Table 2), and between SLIP FreeSurfer and SLIP SynthSeg (median phenotype trajectory correlation, r=0.964; Table 2).

**Table 2.** Brain developmental milestones estimated for the clinical scans with limited imaging pathology (SLIP) using generalized additive models for location, shape and scale (GAMLSS). Models based on imaging phenotypes derived from FreeSurfer (FS) and SynthSeg (SS) image processing pipelines are compared to aggregated research data from the Lifespan Brain Chart Consortium (LBCC). GMV, gray matter volume; WMV, white matter volume; sGMV, subcortical gray matter volume; CSF, ventricle volume; TCV, total cerebrum volume.

| Phenotype | SLIP (FS) Age at Peak (years) | SLIP (SS) Age at Peak (years) | Age-Limited LBCC Age at Peak (years) | SLIP (FS) vs. Age-Limited LBCC 50th Centile (Pearson's r) | SLIP (SS) vs. Age-Limited LBCC 50th Centile (Pearson's r) | SLIP (FS) vs. SLIP (SS) (Pearson's r) |
| --- | --- | --- | --- | --- | --- | --- |
| GMV | 6.06 | 7.35 | 6.55 | 0.999 | 0.999 | 0.954 |
| WMV | 20.1 | 20.1 | 22.1 | 0.990 | 0.997 | 0.964 |
| sGMV | 12.6 | 14.4 | 16.0 | 0.997 | 0.955 | 0.891 |
| CSF | 22.1 | 22.1 | 22.1 | 0.745 | 0.600 | 0.975 |
| TCV | 9.71 | 10.3 | 22.1 | 0.997 | 0.998 | 0.980 |

**Figure 4.**
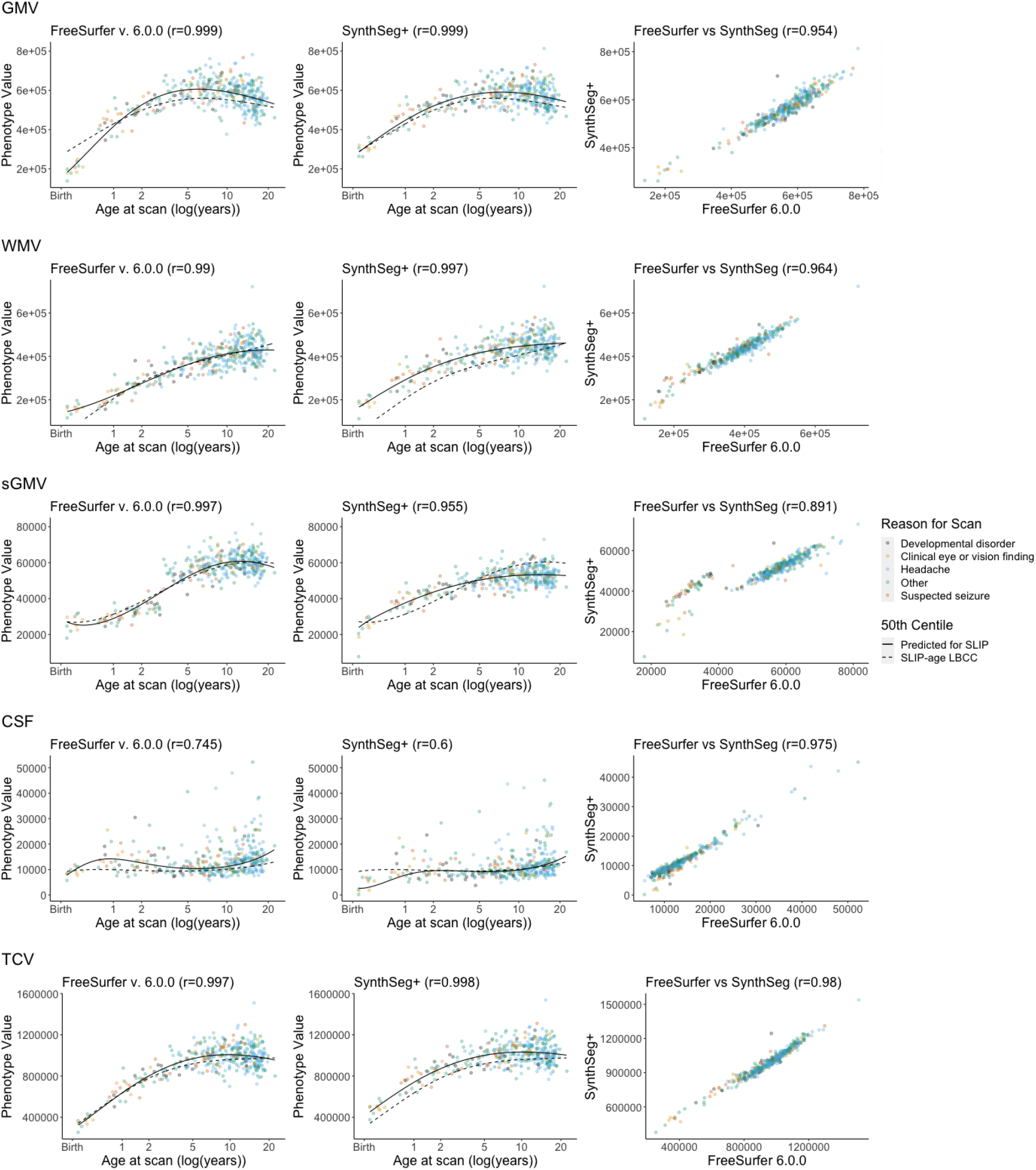
Growth trajectories of the five global imaging phenotypes compared across datasets and image processing pipelines. The clinical scans with limited imaging pathology (SLIP) were processed with FreeSurfer and SynthSeg+, and compared with the models generated from SLIP-aged Lifespan Brain Chart Consortium (LBCC) data. The first two columns display SLIP global imaging phenotypes, the 50th centile line from growth charts of that phenotype estimated with GAMLSS (solid line), and the 50th centile line estimated from the SLIP-age limited LBCC growth charts (dashed line). The third column directly compares the imaging phenotypes derived from FreeSurfer and SynthSeg. GMV, gray matter volume; WMV, white matter volume; sGMV, subcortical gray matter volume; CSF, ventricle volume; TCV, total cerebrum volume.

Despite the high degree of overall similarity, we did observe differences between SLIP and LBCC data: for instance, WMV was observed to peak > 1 year earlier in SLIP compared to LBCC (Table 2). Arguably, differences due to preprocessing pipeline outweighed differences attributable to the dataset: a clear age-related discontinuity was observed for data processed with Infant FS in measurements of sGMV, which has been reported previously (Bethlehem, Seidlitz, and White et al. 2022) (row 3, Figure 4). In addition, measurements of ventricular volume differed significantly between FS and SS, though this difference was largely mitigated when considering centiles benchmarked to GAMLSS models (correlation between ventricular volume centiles in the same individuals r=0.975).

At the resolution of specific cortical regions, strikingly similar maturational trajectories were observed between SLIP and LBCC brain charts via the pattern of interregional differences in the age at peak volume. In both datasets, a clear maturational gradient was observed from early maturation in sensorimotor cortex to late maturation in association cortex (Figure 5). Interregional differences in the age at peak regional volume were significantly correlated between SLIP and LBCC brain charts (Spearman’s r=0.697, P_spin_ < 0.0006) (Alexander-Bloch et al 2018). A general trend was observed such that smaller cortical regions showed greater deviation between SLIP and LBCC brain charts (Figure 5).

**Figure 5.**
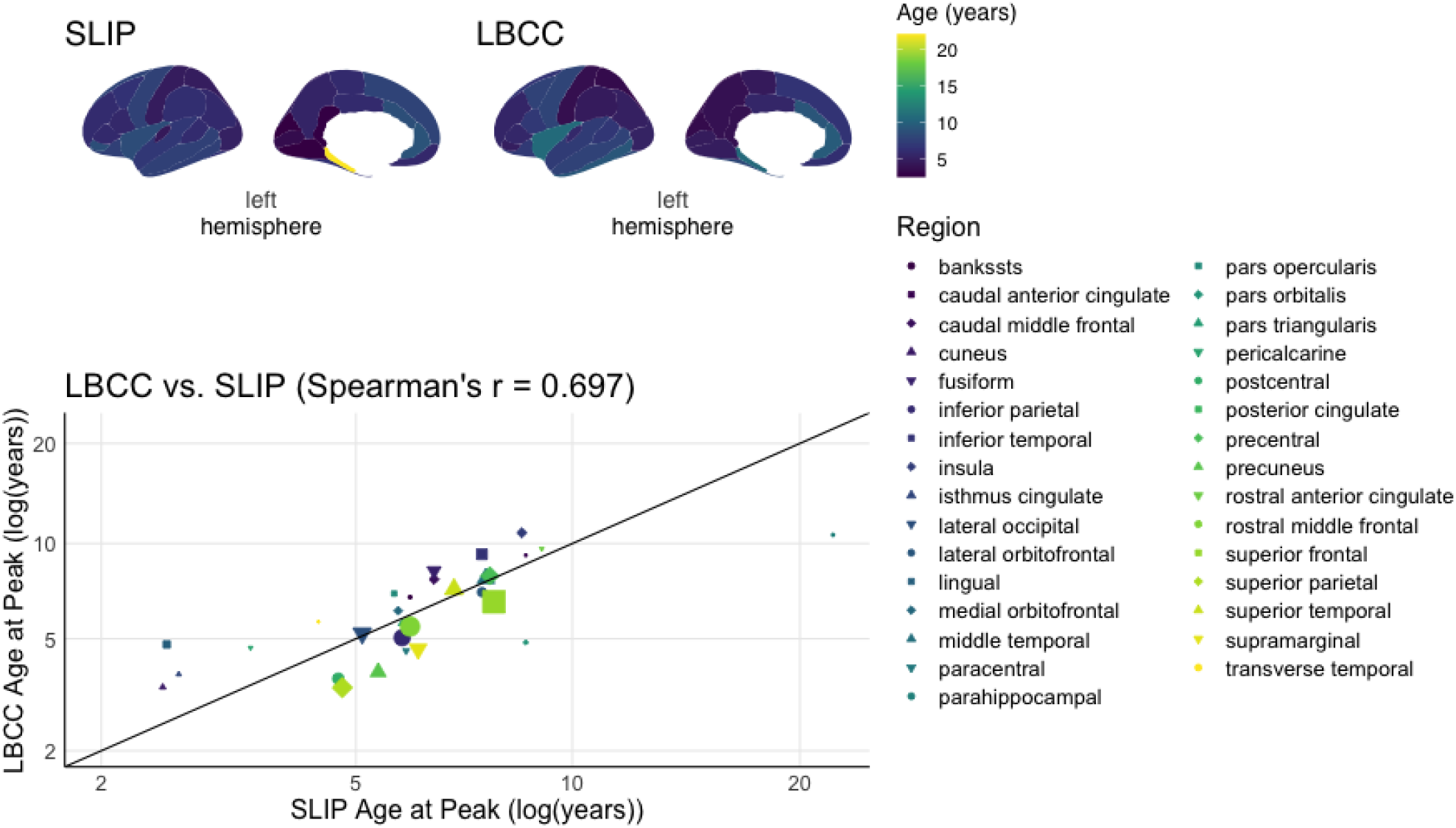
Comparison of regional brain development modeled in clinical controls (SLIP) and research controls (LBCC). In the top panel, age at peak regional volume for cortical regions is illustrated on the cortical surface using the sulcal-based Desikan-Killiany parcellation, with values averaged across left and right hemisphere regions. In the bottom panel, a scatterplot illustrates the correlation between age at peak regional volume in clinical and research controls (Spearman’s r=0.697, P_spin_ < 0.0006). In the scatterplot, the size of each point is proportional to the average size of the brain region it represents.

**Figure 6.**
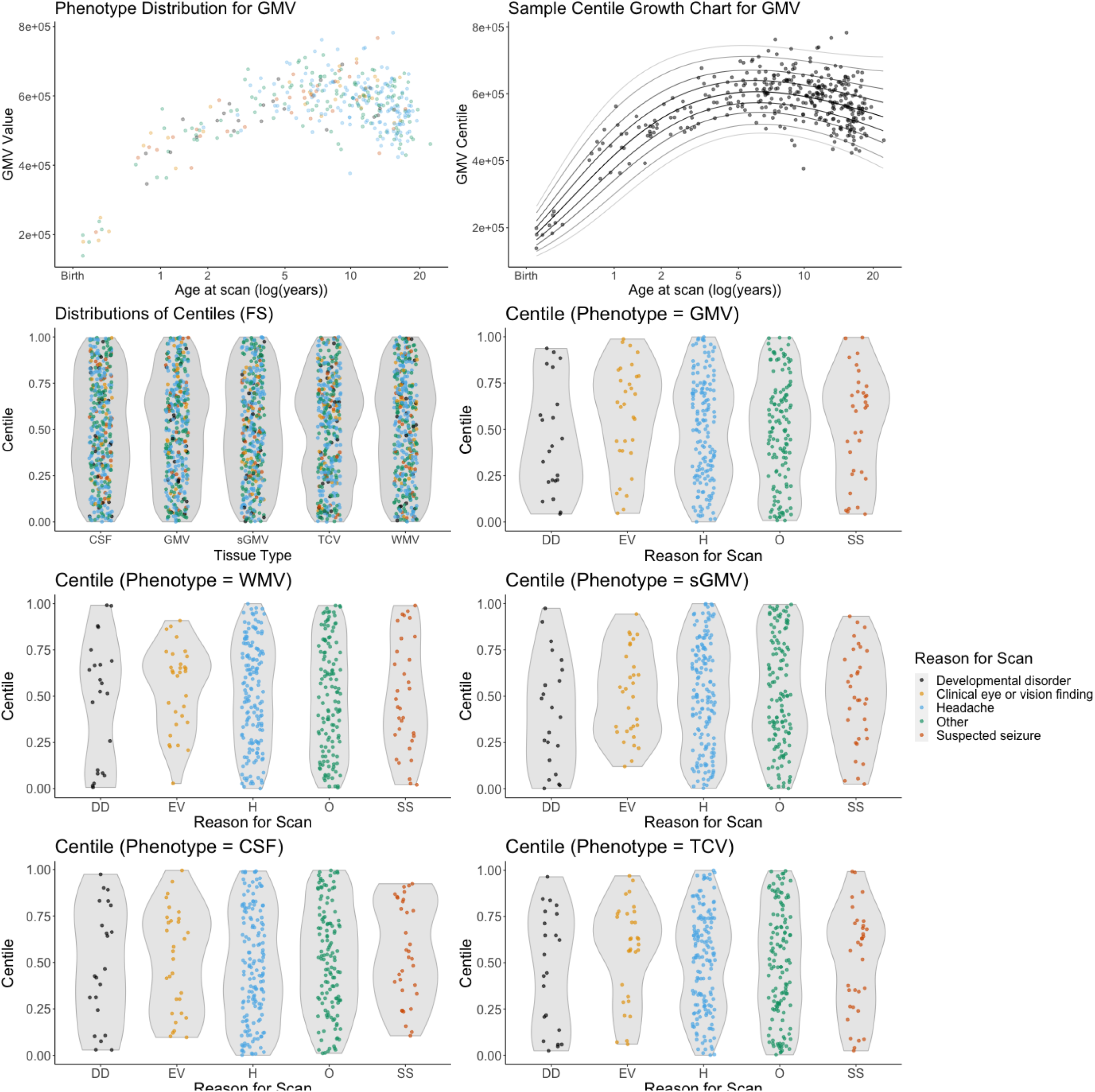
The distribution of phenotype centiles calculated using the GAMLSS models for the global FreeSurfer imaging phenotypes. There was no statistical difference in the centile distributions for each phenotype based on the clinical indication for the scan (developmental disorder, DD; clinical eye or vision finding, EV; headache, H; other, O; suspected seizure, SS). Phenotypes are GMV, gray matter volume; WMV, white matter volume; sGMV, subcortical gray matter volume; CSF, ventricle volume; TCV, total cerebrum volume.

### No evidence of bias by clinical indication for scan

For clinical brain charts to be interpretable as a reference norm, the resulting centiles should not be biased by the clinical indication for patient scans contributing to the reference. We examined the centile distributions for the most frequently occurring reasons for scan in the SLIP cohort, which included the presence of a developmental disorder, clinical eye or vision findings, headaches, and suspected seizures. Critically, we did not find evidence of significantly different outcome variables based on clinical indications between scans (ANOVA P > 0.05 in all cases).

## Discussion

We generated clinical brain growth charts to address the challenge of utilizing existing clinically-acquired data for quantitative brain MRI research. We developed and implemented a curation process to identify clinical MRIs without significant imaging pathology, then performed rigorous quality control and processed them with neuroimaging research pipelines. Following a methodology recommended by the World Health Organization for pediatric growth charts, we used a principled distributional regression technique to model growth charts for global and regional imaging phenotypes (Borghi et al 2006). We compared the scans with limited imaging pathology (SLIP) to one of the largest aggregated brain MRI research datasets (LBCC) and found a high degree of convergence between growth charts derived from each dataset. These results address a longstanding need in brain MRI research by suggesting the utility of clinical brain growth charts as suitable benchmarks for developmental norms.

Our findings are consistent with our prediction that the neuroanatomy of patients who receive negative clinical brain MRIs does not in general diverge from that of research participants recruited as healthy controls. The most common indication for scan in the SLIP dataset was headache: the lifelong prevalence of headache is reported at >95% in the general population, and most scans for headache without focal neurological symptoms are unremarkable (Jordan and Flanders 2020). Similarly, research case-control studies of quantitative anatomical MRI phenotypes in headache including migraine have generally been inconclusive (Sheng et al 2021; Henn et al 2022).

The present work suggests multiple areas for future investigation. Metrics of individual-level deviation from brain charts can be combined with other information available from the electronic health record such as clinical genetics to investigate altered patterns of brain development in subgroups of patients with shared genetic variants. While our effort to compare clinical- and research-controls benefited from the radiology department’s use of a high-quality isotropic T1-weighted MRI sequence for routine brain scans, SynthSeg+ is expected to generalize well to lower quality scans and diverse clinical scan protocols (Billot et al. 2022). Greater inclusion of scans would increase the sample size incorporated into our reference charts in early developmental and adult populations. Finally, although short-term utility is limited to the research domain, it is conceivable that contemporaneous, growth chart benchmarks of quantitative imaging features could prove to be clinically useful in certain settings.

## Data Availability

All data produced in the present study are currently unable to be shared but may be in the future.

## Acknowledgements

The views expressed are those of the author(s) and not necessarily those of the NIHR or the Department of Health and Social Care. We thank Jisoo Kang, Nadia Ngom, Sreya Subramanian, and Caleb Schmitt for contributing to the review of signed radiology reports. Additional support was provided by the CHOP-Penn Lifespan Brain Institute. The live list of authors included in the Lifespan Brain Chart Consortium can be found at https://github.com/brainchart/Lifespan.

## Abbreviations

SLIP: Scans with Limited Imaging Pathology
LBCC: Lifespan Brain Chart Consortium
GMV: cortical Gray Matter Volume
WMV: White Matter Volume
sGMV: Subcortical Gray Matter Volume
CSF: ventricular volume (Cerebrospinal Fluid)
TCV: Total Cerebrum Volume

## Supplementary Materials

### Harmonized MPRAGE T1-weighted Sequence

The radiology department implemented a standardized high resolution T1 weighted scanning protocol included in every clinical scan session starting in 2008. The sequence is a sagittal MPRAGE with voxel resolution of 0.9mm × 0.9mm and slice thickness of 0.9mm with ranges of TE = 1900-2050ms, TR = 2.45-2.55ms, and TI = 900-1050ms based on scanner bore size and gradient strengths. The dimensions of each scan are 256 × 256 voxels per slice and 192 slices, although the technologist had the liberty of decreasing the number of slices in the sagittal plane as needed to avoid spending gradient time scanning non-tissue (air) depending on patient head size. The estimated time to obtain a single scan using this protocol is approximately 4:26 minutes for the full acquisition. The reason for this implementation was to achieve a uniform isotropic acquisition with adequate gray-white differentiation across clinical imaging studies that would serve the evaluation of a wide range of pathologies and could be easily reformatted into other planes as needed.

### Data Curation

First, an SQL query was performed to obtain radiology reports of brain MRI scans taken between 1992 and 2021 (N > 120,000 reports). Each radiology report contained both findings and overall impressions and was signed by a licensed neuroradiologist. A subset of the total reports were manually examined to identify scans where no significant imaging pathology was reported. The positive findings were described in terms of the tissue abnormalities, their radiographic properties, and their location while negative findings were explicitly described a lack of tissue abnormalities. Anonymous exemplar reports in Supplementary Table 1 demonstrate positive radiology findings and negative radiology findings, which were excluded and included in the SLIP dataset, respectively. To date, 10,116 reports have been manually examined of which 3,987 have been identified as SLIP.

A list of 1,013 scan sessions from the identified SLIP reports (N=3,987) were compiled into a scan request. The request for these scan sessions was submitted to the radiology department by an honest broker (a person who is authorized to view clinical data with the purpose of removing identifying information on behalf of another party) using biomedical informatics resources hosted at the hospital. The honest broker obtained anonymized scans for 731 sessions from the *p*icture *a*rchiving and *c*ommunication *s*ystem (PACS) server used by the radiology department to store, organize, and communicate about medical imaging data. The 30% data loss between the number of requested scans and the number of delivered scans results from the missing sessions not being available on the PACS server either because the scan was from an external site or because the scan was not migrated to digital storage when digital scan storage first became available.

The scans were converted from DICOM format to NIFTI format using the tool *heudiconv* (version 0.9.0) (Halchenko et al 2020) and organized according to the Brain Imaging Data Structure (BIDS) standard (Gorgolowski et al 2016) using the *CuBIDS* tool developed by Covitz et al (Covitz et al 2022). *CuBIDs* was then used to filter the data based on parameters in the metadata. Initial filtering to remove scans with large voxels and large interslice distances produced a set of 1,266 high resolution MPR scans. Of these scans, 22 were removed due to missing metadata (N=1,244) and 72 were removed because they had been derived at the scanner workstation (N=1,172). The first high resolution (1×1×1 mm or smaller voxels) non-contrast MPRAGE scan with more than 60 slices was designated as the representative scan for each 3T session. The identification of the representative scans was greatly aided by the radiology department’s standardized ∼1mm isotropic MPRAGE T1-weighted sequence (see above).

### Manual Quality Assessment

The representative scans underwent manual and automated image quality assessment (Rosen et al 2018; Bedford et al 2022). The scans were stratified into batches of at most 80 scans based on age group (0-2 years, 2-5 years, 5-10 years, 10+ years). For each scan, three slices from each dimension (9 slices per scan) were pseudorandomly selected from a range of 20 slices centered around the origin (in MNI space) and midpoints in each cardinal direction. The slices were saved as .PNG files and two independent raters (interrater agreement across scans of 0.917) evaluated image quality of each using a Jupyter notebook accessed via a HIPAA-compliant server. The images were rated as being poor quality (grade of 0), acceptable quality (grade of 1), or highest quality (grade of 2). The grades of the images were averaged across the raters and scans with averages of 1.0 and higher were used in the statistical analysis.

### Processing

Two segmentation pipelines were used to extract imaging phenotypes: a FreeSurfer based pipeline and a SynthSeg based pipeline.

The FreeSurfer pipeline used several FSL tools to preprocess the scans: it first reoriented the scan to the MNI152 atlas (Fonov et al 2011) using *fslreorient2std* (Jenkinson et al 2012), removed the facial features using *mri_deface*, and finally performed a rigid AC/PC alignment to MNI152 using *flirt*. The preprocessed scans underwent image segmentation using the full *recon-all* pipeline from either FreeSurfer 6.0.0 (Fischl et al 2004) for patients over the age of 3 years or Infant Freesurfer 6.0.0 for patients under the age of 3 years (Zöllei et al 2020). The SynthSeg pipeline performed brain and subcortical structure segmentation using SynthSeg+ (Billot et al. 2021). SynthSeg+ is an emerging deep learning based algorithm designed to perform automated brain segmentation on scans FreeSurfer might find difficult to work with due to resolution issues or patient motion. We used the pretrained models available on GitHub (https://github.com/BBillot/SynthSeg).

Each pipeline produced quantitative volumetric measurements subsequently referred to as “imaging phenotypes” that were used along with demographic and quality assessment information for each scan in downstream analyses. A description of the five primary volumetric imaging phenotypes and how they were calculated for each pipeline can be found in Supplemental Table 4. Additionally, the FreeSurfer pipeline produced 34 regional cortical volumes quantified using the sulcal-based Desikan-Killiany parcellation (Desikan et al 2006). An additional metric-based quality check was performed to ensure neither pipeline produced egregiously poor segmentations. The relative difference between corresponding imaging phenotypes from each pipeline was calculated. Two scans with relative differences close to 2 across all five primary volumetric imaging phenotypes were excluded. These scans were for patients under the age of 2 weeks post-birth and contained more motion in their image volumes than seen in the manual quality assessment.

### Statistical Analysis

We sought to compare clinical brain growth charts to previously published, publicly available research brain growth charts derived from the LBCC reference dataset of 123,984 MRIs across more than 100 primary studies (www.brainchart.io) (Bethlehem, Seidlitz, and White et al. 2022). As detailed previously, brain growth charts were constructed using generalized additive models for location, scale, and shape (GAMLSS): a distributional regression approach that allows for modeling the mean, variance, and higher-order statistical moments in terms of flexible nonlinear associations using covariates of interest (Stasinopoulos et al 2017). For a more accurate comparison of the five global volumetric imaging phenotypes in this work, a subset of the LBCC data processed either using FreeSurfer 6.0.0 or Infant FreeSurfer was used as the basis for similarly constructed SLIP-age limited LBCC GAMLSS growth charts.

Prior to growth chart modeling, we used ComBat, a batch correction approach adapted from statistical genomics, to harmonize the imaging phenotypes across scanners (Johnson and Li 2007; Fortin et al 2017; Pomponio et al 2019). Our previous work suggested that harmonization was comparable across using either ComBat or GAMLSS (including site as a random effect in GAMLSS models). In the present paper we elected to use ComBat to harmonize between scanners, which allowed the whole SLIP cohort to be treated as a single study in comparison with the Lifespan models. Specifically, we used the Python implementation of *neuroHarmonize* (version 2.1.0) (https://github.com/rpomponio/neuroHarmonize) (Pomponio et al, 2019). ComBat was used to control for the effect of MR scanners while preserving the effect of other model covariates, notably age at scan, patient sex, and clinical indication for scan.

GAMLSS was used to fit growth charts of SLIP data following a similar protocol as LBCC. A generalized gamma distribution was used to link each imaging phenotype to predictor variables using the GAMLSS package in R (version 5.4-10) (Stasinopoulos et al 2017). The predictor variables in these models included age and sex as reported in the electronic health record. For the FreeSurfer-based analysis, the Euler number, a robust marker of image quality, was also included (Rosen et al. 2018). As detailed previously for LBCC growth charts, the mean and variance of each GAMLSS was modeled by a non-linear age effect using third-order fractional polynomials, while an intercept term alone was employed for model skewness allowing for shared skewness across the age range.

**Supplemental Table 1.**
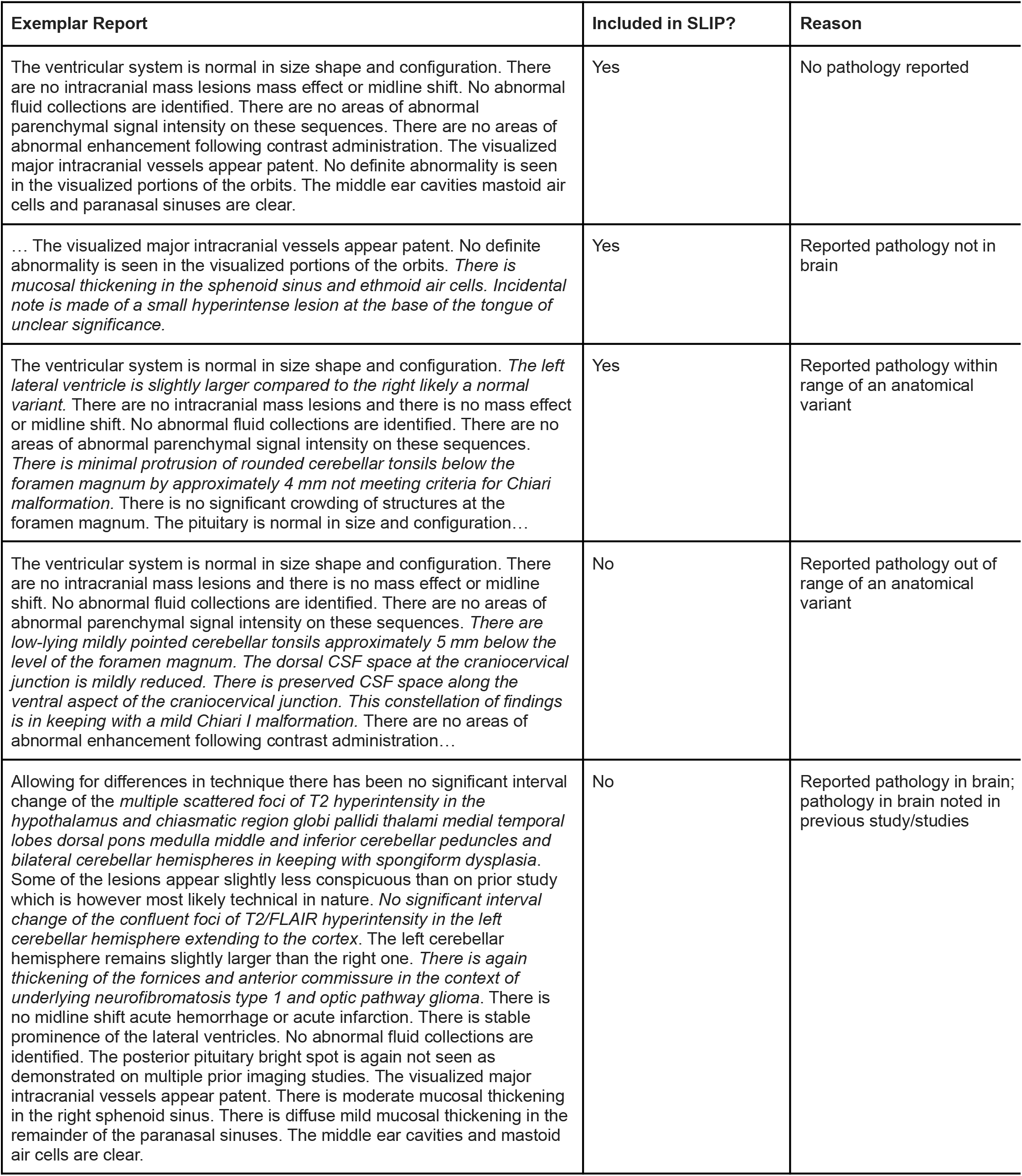
Anonymous examples of radiology reports included in and excluded from the SLIP dataset. Exemplar reports have been edited for brevity while including context supporting inclusion/exclusion of the data from the SLIP cohort. Sentences pertaining to pathology have been italicized.

| Exemplar Report | Included in SLIP? | Reason |
| --- | --- | --- |
| The ventricular system is normal in size shape and configuration. There are no intracranial mass lesions mass effect or midline shift. No abnormal fluid collections are identified. There are no areas of abnormal parenchymal signal intensity on these sequences. There are no areas of abnormal enhancement following contrast administration. The visualized major intracranial vessels appear patent. No definite abnormality is seen in the visualized portions of the orbits. The middle ear cavities mastoid air cells and paranasal sinuses are clear. | Yes | No pathology reported |
| ... The visualized major intracranial vessels appear patent. No definite abnormality is seen in the visualized portions of the orbits. *There is mucosal thickening in the sphenoid sinus and ethmoid air cells. Incidental note is made of a small hyperintense lesion at the base of the tongue of unclear significance.* | Yes | Reported pathology not in brain |
| The ventricular system is normal in size shape and configuration. *The left lateral ventricle is slightly larger compared to the right likely a normal variant.* There are no intracranial mass lesions and there is no mass effect or midline shift. No abnormal fluid collections are identified. There are no areas of abnormal parenchymal signal intensity on these sequences. *There is minimal protrusion of rounded cerebellar tonsils below the foramen magnum by approximately 4 mm not meeting criteria for Chiari malformation.* There is no significant crowding of structures at the foramen magnum. The pituitary is normal in size and configuration... | Yes | Reported pathology within range of an anatomical variant |
| The ventricular system is normal in size shape and configuration. There are no intracranial mass lesions and there is no mass effect or midline shift. No abnormal fluid collections are identified. There are no areas of abnormal parenchymal signal intensity on these sequences. *There are low-lying mildly pointed cerebellar tonsils approximately 5 mm below the level of the foramen magnum. The dorsal CSF space at the craniocervical junction is mildly reduced. There is preserved CSF space along the ventral aspect of the craniocervical junction. This constellation of findings is in keeping with a mild Chiari I malformation.* There are no areas of abnormal enhancement following contrast administration... | No | Reported pathology out of range of an anatomical variant |
| Allowing for differences in technique there has been no significant interval change of the *multiple scattered foci of T2 hyperintensity in the hypothalamus and chiasmatic region globi pallidi thalami medial temporal lobes dorsal pons medulla middle and inferior cerebellar peduncles and bilateral cerebellar hemispheres in keeping with spongiform dysplasia.* Some of the lesions appear slightly less conspicuous than on prior study which is however most likely technical in nature. *No significant interval change of the confluent foci of T2/FLAIR hyperintensity in the left cerebellar hemisphere extending to the cortex.* The left cerebellar hemisphere remains slightly larger than the right one. *There is again thickening of the fornices and anterior commissure in the context of underlying neurofibromatosis type 1 and optic pathway glioma.* There is no midline shift acute hemorrhage or acute infarction. There is stable prominence of the lateral ventricles. No abnormal fluid collections are identified. The posterior pituitary bright spot is again not seen as demonstrated on multiple prior imaging studies. The visualized major intracranial vessels appear patent. There is moderate mucosal thickening in the right sphenoid sinus. There is diffuse mild mucosal thickening in the remainder of the paranasal sinuses. The middle ear cavities and mastoid air cells are clear. | No | Reported pathology in brain; pathology in brain noted in previous study/studies |

**Supplemental Table 2.** Demographics of the LBCC subset of the same age range as the SLIP data used to generate the median centiles in the global imaging phenotype trajectories. Participants in these studies older than 22 years were excluded from the comparison with the SLIP data.

| **Study** | **Age Range (Median) in Years** | **Total** | **Female** | **Male** |
| --- | --- | --- | --- | --- |
| *Total* | 0.42 - 22 (14) | 8346 | 3399 | 4947 |
| ABCD | 9 - 10.9 (9.7) | 376 | 115 | 221 |
| ABIDE1 | 6.5 - 22 (13.6) | 859 | 136 | 723 |
| ABIDE2 | 5.1 - 22 (11.5) | 647 | 139 | 508 |
| ADHD200 | 7.2 - 21.7 (11.2) | 809 | 294 | 515 |
| AOBA | 11.2 - 21.2 (19.2) | 132 | 39 | 93 |
| AOMIC 1000 | 19 - 22 (21) | 362 | 187 | 175 |
| ASRB | 18 - 22 (21) | 37 | 21 | 16 |
| BHRCS | 5.8 - 21 (10.1) | 717 | 314 | 403 |
| BSNIP | 8.9 - 22 (18.2) | 147 | 71 | 76 |
| CAMFT | 12.5 - 16.4 (14.9) | 68 | 35 | 33 |
| cVEDA | 4.7 - 22 (14.6) | 1162 | 482 | 680 |
| EMBARC | 17.3 - 22 (19.2) | 41 | 30 | 11 |
| Female ASD | 6.1 - 17.2 (11.6) | 358 | 180 | 178 |
| IBIS | 0.42 - 2.2 (0.6) | 394 | 148 | 246 |
| ICBM | 17.2 - 21.2 (19.2) | 149 | 82 | 67 |
| IMAGEN | 12 - 22 (14) | 1792 | 912 | 880 |
| LA5c | 21 - 22 (21) | 29 | 18 | 11 |
| MCIC | 17.2 - 21.2 (19.2) | 47 | 14 | 33 |
| Narratives | 2 - 22 (20) | 135 | 75 | 60 |
| SALD | 19 - 22 (21) | 40 | 28 | 14 |
| STRIVE | 18.3 - 22 (20.7) | 35 | 35 | 0 |
| WAYNE | 18 - 22 (19.5) | 10 | 6 | 4 |

**Supplemental Table 3.** Specifications and number of scans for each scanner. Scanner ID assigned based on number of scans.

| Scanner ID | Number of Scans | Manufacturer | Model Name | Software Versions |
| --- | --- | --- | --- | --- |
| Scanner 1 | 155 | Siemens | TrioTim | Syngo MR 2006T 4VB12T |
| Scanner 2 | 84 | Siemens | TrioTim | Syngo MR B15 |
| Scanner 3 | 84 | Siemens | Verio | Syngo MR B15V |
| Scanner 4 | 14 | Siemens | Trio | Syngo MR 2004A 4VA25A |
| Scanner 5 | 13 | Siemens | Verio | Syngo MR B15V |
| Scanner 6 | 11 | Siemens | Trio | Syngo MR 2004A 4VA25A |
| Scanner 7 | 6 | Siemens | Skyra | Syngo MR E11 |
| Scanner 8 | 2 | Siemens | Skyra | Syngo MR D13C |
| Scanner 9 | 3 | Siemens | Prisma Fit | Syngo MR D13D |

**Supplemental Table 4.** A description of how each of the primary five volumetric imaging phenotypes were calculated from the FreeSurfer and SynthSeg+ outputs.

| Imaging Phenotype | FreeSurfer | SynthSeg+ |
| --- | --- | --- |
| GMV (Gray Matter Volume) | CortexVol | left cerebral cortex + right cerebral cortex |
| WMV (White Matter Volume) | CerebralWhiteMatterVol | left cerebral white matter + right cerebral white matter |
| sGMV (Subcortical Gray Matter Volume) | SubCortGrayVol | left thalamus + left caudate + left putamen + left pallidum + left hippocampus + left amygdala + left accumbens area + right thalamus + right caudate + right putamen + right pallidum + right hippocampus + right amygdala + right accumbens area |
| CSF (Ventricular Volume) | BrainSegVol - BrainSegVolNotVent | left lateral ventricle + right lateral ventricle + left inferior lateral ventricle + right inferior lateral ventricle + 3rd ventricle + 4th ventricle |
| TCV (Total Cerebrum Volume) | GMV + WMV | GMV + WMV |

**Supplementary Figure 1.**
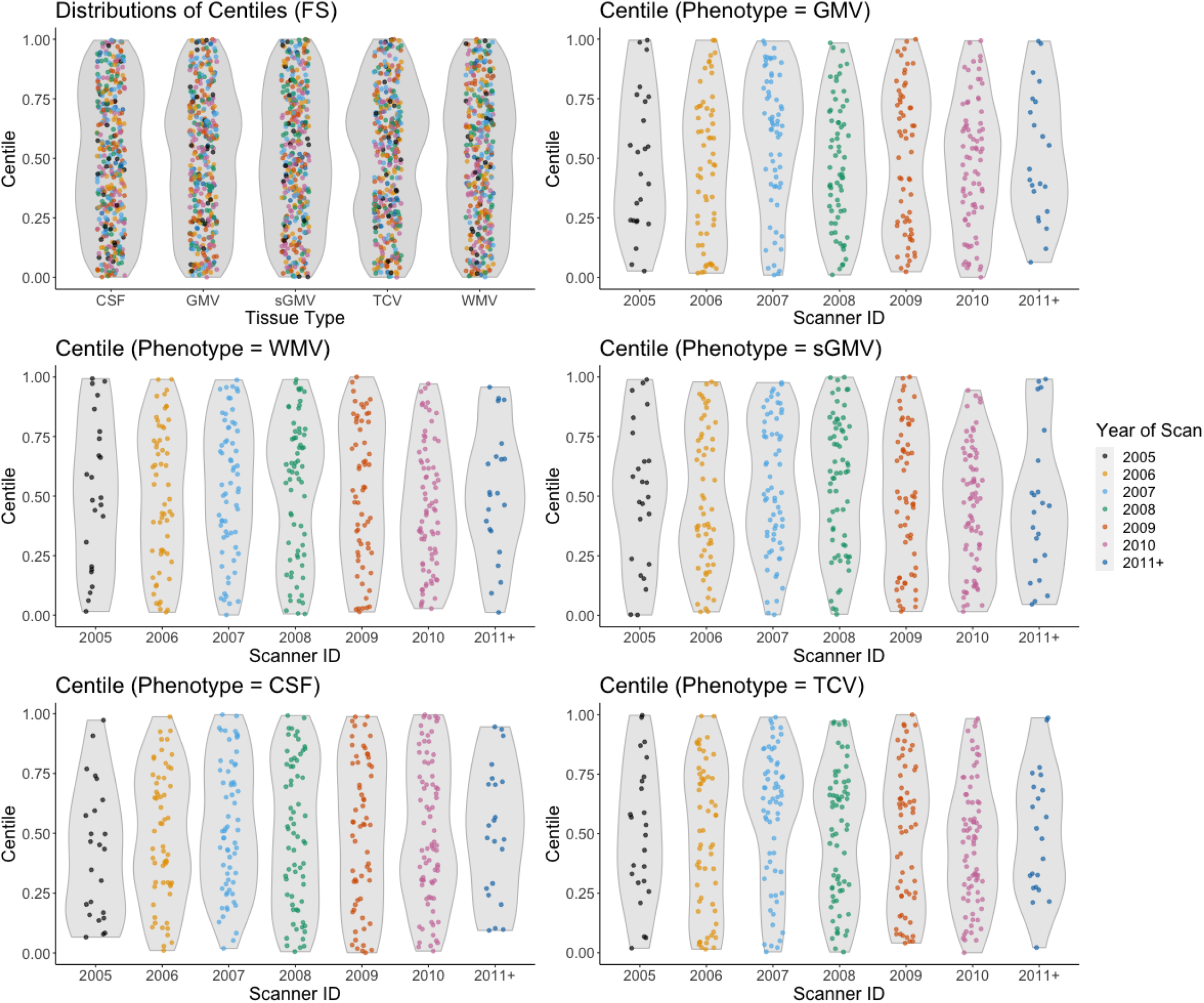
The distribution of phenotype centiles calculated using the GAMLSS models for the global FreeSurfer imaging phenotypes displayed in relation to the year of each scan.

**Supplementary Figure 2.**
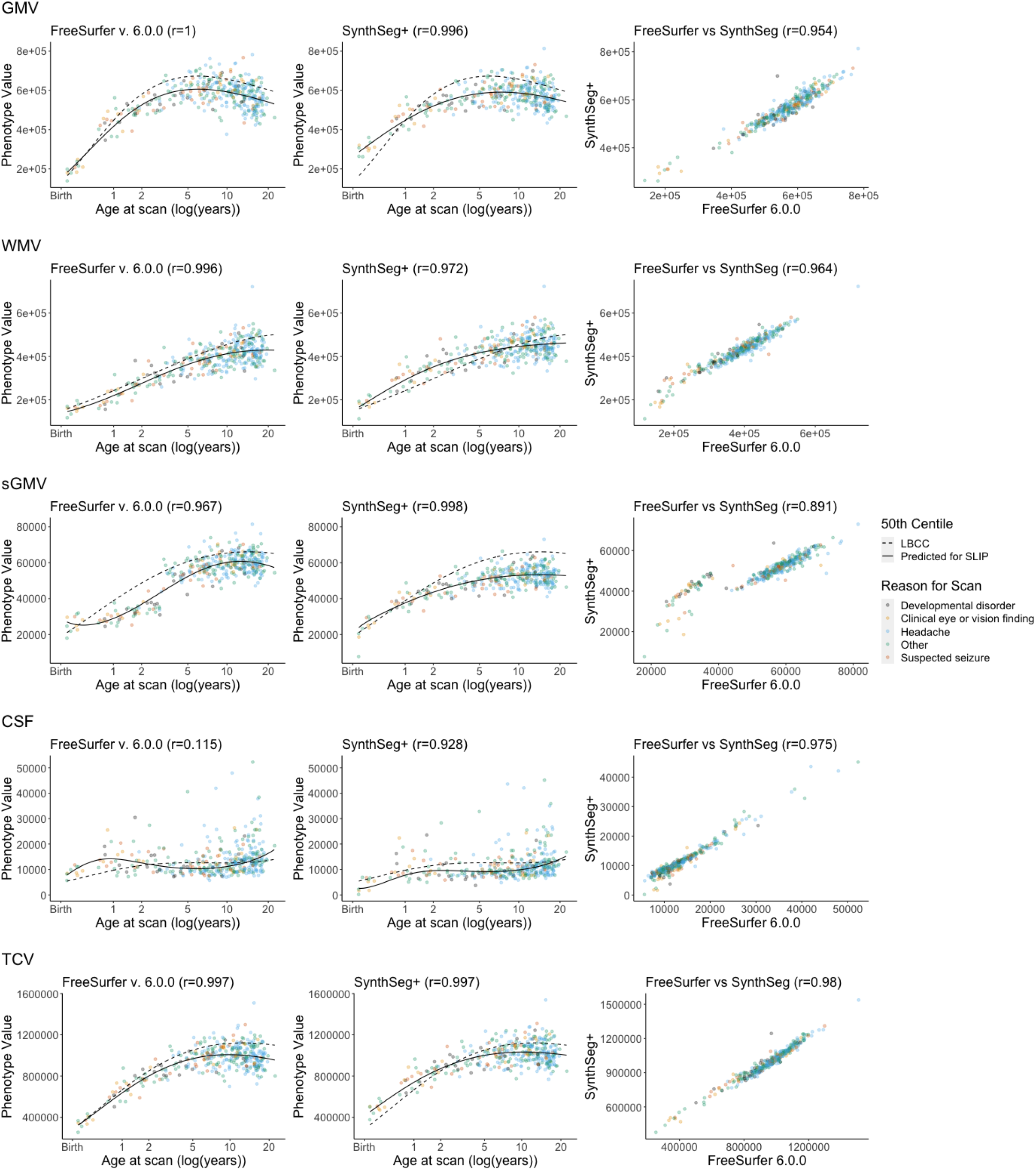
Growth trajectories of the five global imaging phenotypes compared across datasets and image processing pipelines compared to the full lifespan growth chart models publicly available from the Lifespan Brain Chart Consortium (LBCC). The clinical scans with limited imaging pathology (SLIP) were processed with FreeSurfer and SynthSeg. For each imaging phenotype, the first column displays the FS phenotype, the 50th centile line from growth charts of that phenotype estimated with GAMLSS (solid line), and the 50th centile line from the full lifespan LBCC growth charts also estimated with GAMLSS (dashed line). The second column displays the SS phenotype, the 50th centile line from growth charts of that phenotype estimated with GAMLSS (solid line), and the 50th centile line from the full lifespan LBCC growth charts (dashed line). GMV, gray matter volume; WMV, white matter volume; sGMV, subcortical gray matter volume; CSF, ventricle volume; TCV, total cerebrum volume.

## References

1. Yaroslav Halchenko; Mathias Goncalves; Matteo Visconti di Oleggio Castello; Satrajit Ghosh; Taylor Salo;Michael Hanke; Dae; Pablo Velasco; James Kent; Matthew Brett; Inge Amlien; Chris Gorgolewski; Darren Christopher Lukas; Chris Markiewicz; Steven Tilley; Jakub Kaczmarzyk; Joerg Stadler; Sin Kim; Ari Kahn; Benjamin Poldrack; Bruno Melo; Henry Braun; John Pellman; Daniel Lurie; john lee; Adina Wagner; Franklin Feingold; Johan Carlin; Kalle Samuels; Michał Szczepanik. A flexible DICOM converter for organizing brain imaging data into structured directory layouts. December 23, 2020. DOI: 10.5281/zenodo.4390433.

2. Gorgolewski, K.J., Auer, T., Calhoun, V.D., Craddock, R.C., Das, S., Duff, E.P., Flandin, G., Ghosh, S.S., Glatard, T., Halchenko, Y.O. and Handwerker, D.A., 2016. The brain imaging data structure, a format for organizing and describing outputs of neuroimaging experiments. Scientific data, 3(1), pp.1–9.

3. Covitz S, Tapera TM, Adebimpe A, Alexander-Bloch A, Bertolo MA, Feczko E, Franco AR, Gur RE, Gur RC, Hendrickson T, Houghton A, Mehta K, Murtha K, Perrone AJ, Robert-Fitzgerald T, Schabdach JM, Shinohara RT, Vogel JW, Zhao C, Fair DA, Milham MP, Cieslak M, Satterthwaite TD (2022): Curation of BIDS (CuBIDS): a workflow and software package for streamlining reproducible curation of large BIDS datasets. NeuroImage 263, 11960 (2022). https://doi.org/10.1016/j.neuroimage.2022.119609

4. W. Evan Johnson and Cheng Li, Adjusting batch effects in microarray expression data using empirical Bayes methods. Biostatistics, 8(1):118–127, 2007. https://doi.org/10.1093/biostatistics/kxj037.

5. Fortin, J. P., N. Cullen, Y. I. Sheline, W. D. Taylor, I. Aselcioglu, P. A. Cook, P. Adams, C. Cooper, M. Fava, P. J. McGrath, M. McInnis, M. L. Phillips, M. H. Trivedi, M. M. Weissman and R. T. Shinohara (2017). “Harmonization of cortical thickness measurements across scanners and sites.” Neuroimage 167: 104–120. https://doi.org/10.1016/j.neuroimage.2017.11.024.

6. Pomponio, R., Shou, H., Davatzikos, C., et al., (2019). “Harmonization of large MRI datasets for the analysis of brain imaging patterns throughout the lifespan.” Neuroimage 208. https://doi.org/10.1016/j.neuroimage.2019.116450.

7. VS Fonov, AC Evans, K Botteron, CR Almli, RC McKinstry, DL Collins and BDCG, Unbiased average age-appropriate atlases for pediatric studies, NeuroImage, Volume 54, Issue 1, January 2011, ISSN 1053–8119, DOI: 10.1016/j.neuroimage.2010.07.033

8. M. Jenkinson, C.F. Beckmann, T.E. Behrens, M.W. Woolrich, S.M. Smith. FSL. NeuroImage, 62:782–90, 2012

9. Bruce Fischl, Andre van der Kouwe, Christophe Destrieux, Eric Halgren, Florent Segonne, David H. Salat, Evelina Busa, Larry J. Seidman, Jill Goldstein, David Kennedy, Verne Caviness, Nikos Makris, Bruce Rosen, and Anders M. Dale. Automatically Parcellating the Human Cerebral Cortex. Cerebral Cortex January 2004; 14:11–22.

10. Rosen, A.F., Roalf, D.R., Ruparel, K., Blake, J., Seelaus, K., Villa, L.P., Ciric, R., Cook, P.A., Davatzikos, C., Elliott, M.A. and de La Garza, A.G., 2018. Quantitative assessment of structural image quality. Neuroimage, 169, pp.407–418.

11. Bedford SA, Ortiz-Rosa A, Schabdach JM, Costantino M, Tullo S, Piercy T, Lai M-C, Lombardo M V, Di Martino A, Devenyi GA, Chakravarty MM, Alexander-Bloch AF, Seidlitz J, Baron-Cohen S, Bethlehem RAI (2022): The impact of quality control on cortical morphometry comparisons in autism. medRxiv:2022.12.05.22283091. http://medrxiv.org/content/early/2022/12/06/2022.12.05.22283091.abstract.

12. Zöllei, L, Iglesias, JE, Ou, Y, Grant, PR, Fischl, B. Infant FreeSurfer: An automated segmentation and surface extraction pipeline for T1-weighted neuroimaging data of infants 0-2 years, NeuroImage, 2020, 116946, https://doi.org/10.1016/j.neuroimage.2020.116946.

13. Benjamin Billot, Colin Magdamo, You Cheng, Steven E. Arnold, Sudeshna Das, J. E. Iglesias. Robust machine learning segmentation for large-scale analysis of heterogeneous clinical brain MRI datasets. In 2209.02032 (2022)

14. McClure, P., Rho, N., Lee, J.A., Kaczmarzyk, J.R., Zheng, C.Y., Ghosh, S.S., Nielson, D.M., Thomas, A.G., Bandettini, P. and Pereira, F., 2019. Knowing what you know in brain segmentation using Bayesian deep neural networks. Frontiers in neuroinformatics, 13, p.67.

15. Stasinopoulos, M.D., Rigby, R.A., Heller, G.Z., Voudouris, V. and De Bastiani, F., 2017. Flexible regression and smoothing: using GAMLSS in R. CRC Press.

16. Desikan, R.S., Ségonne, F., Fischl, B., Quinn, B.T., Dickerson, B.C., Blacker, D., Buckner, R.L., Dale, A.M., Maguire, R.P., Hyman, B.T. and Albert, M.S., 2006. An automated labeling system for subdividing the human cerebral cortex on MRI scans into gyral based regions of interest. Neuroimage, 31(3), pp.968–980.

17. Rosen AFG, Roalf DR, Ruparel K, Blake J, Seelaus K, Villa LP, Ciric R, Cook PA, Davatzikos C, Elliott MA, Garcia de La Garza A, Gennatas ED, Quarmley M, Schmitt JE, Shinohara RT, Tisdall MD, Craddock RC, Gur RE, Gur RC, Satterthwaite TD. Quantitative assessment of structural image quality. Neuroimage. 2018 Apr 1;169:407–418. DOI: 10.1016/j.neuroimage.2017.12.059. Epub 2017 Dec 24. PMID: 29278774; PMCID: PMC5856621.

18. Bethlehem RAI, Seidlitz J, White SR, Vogel JW, Anderson KM, Adamson C, Adler S, Alexopoulos GS, Anagnostou E, Areces-Gonzalez A, Astle DE, Auyeung B, Ayub M, Bae J, Ball G, Baron-Cohen S, Beare R, Bedford SA, Benegal V, Beyer F, Blangero J, Blesa Cábez M, Boardman JP, Borzage M, Bosch-Bayard JF, Bourke N, Calhoun VD, Chakravarty MM, Chen C, Chertavian C, Chetelat G, Chong YS, Cole JH, Corvin A, Costantino M, Courchesne E, Crivello F, Cropley VL, Crosbie J, Crossley N, Delarue M, Delorme R, Desrivieres S, Devenyi GA, Di Biase MA, Dolan R, Donald KA, Donohoe G, Dunlop K, Edwards AD, Elison JT, Ellis CT, Elman JA, Eyler L, Fair DA, Feczko E, Fletcher PC, Fonagy P, Franz CE, Galan-Garcia L, Gholipour A, Giedd J, Gilmore JH, Glahn DC, Goodyer IM, Grant PE, Groenewold NA, Gunning FM, Gur RE, Gur RC, Hammill CF, Hansson O, Hedden T, Heinz A, Henson RN, Heuer K, Hoare J, Holla B, Holmes AJ, Holt R, Huang H, Im K, Ipser J, Jack CR, Jackowski AP, Jia T, Johnson KA, Jones PB, Jones DT, Kahn RS, Karlsson H, Karlsson L, Kawashima R, Kelley EA, Kern S, Kim KW, Kitzbichler MG, Kremen WS, Lalonde F, Landeau B, Lee S, Lerch J, Lewis JD, Li J, Liao W, Liston C, Lombardo M V., Lv J, Lynch C, Mallard TT, Marcelis M, Markello RD, Mathias SR, Mazoyer B, McGuire P, Meaney MJ, Mechelli A, Medic N, Misic B, Morgan SE, Mothersill D, Nigg J, Ong MQW, Ortinau C, Ossenkoppele R, Ouyang M, Palaniyappan L, Paly L, Pan PM, Pantelis C, Park MM, Paus T, Pausova Z, Paz-Linares D, Pichet Binette A, Pierce K, Qian X, Qiu J, Qiu A, Raznahan A, Rittman T, Rodrigue A, Rollins CK, Romero-Garcia R, Ronan L, Rosenberg MD, Rowitch DH, Salum GA, Satterthwaite TD, Schaare HL, Schachar RJ, Schultz AP, Schumann G, Schöll M, Sharp D, Shinohara RT, Skoog I, Smyser CD, Sperling RA, Stein DJ, Stolicyn A, Suckling J, Sullivan G, Taki Y, Thyreau B, Toro R, Traut N, Tsvetanov KA, Turk-Browne NB, Tuulari JJ, Tzourio C, Vachon-Presseau É, Valdes-Sosa MJ, Valdes-Sosa PA, Valk SL, van Amelsvoort T, Vandekar SN, Vasung L, Victoria LW, Villeneuve S, Villringer A, Vértes PE, Wagstyl K, Wang YS, Warfield SK, Warrier V, Westman E, Westwater ML, Whalley HC, Witte A V., Yang N, Yeo B, Yun H, Zalesky A, Zar HJ, Zettergren A, Zhou JH, Ziauddeen H, Zugman A, Zuo XN, Rowe C, Frisoni GB, Binette AP, Bullmore ET, Alexander-Bloch AF (2022): Brain charts for the human lifespan. Nature:1–26. https://www.nature.com/articles/s41586-022-04554-y.

19. Aaron F. Alexander-Bloch, Haochang Shou, Siyuan Liu, Theodore D. Satterthwaite, David C. Glahn, Russell T. Shinohara, Simon N. Vandekar, Armin Raznahan. On testing for spatial correspondence between maps of human brain structure and function. NeuroImage. Volume 178, 2018. Pages 540–551. ISSN 1053-8119. https://doi.org/10.1016/j.neuroimage.2018.05.070.

20. Borghi, E., de Onis, M., Garza, C., Van den Broeck, J., Frongillo, E.A., Grummer-Strawn, L., Van Buuren, S., Pan, H., Molinari, L., Martorell, R. and Onyango, A.W., 2006. Construction of the World Health Organization child growth standards: selection of methods for attained growth curves. Statistics in medicine, 25(2), pp.247–265.

21. Jordan, J.E. and Flanders, A.E., 2020. Headache and neuroimaging: why we continue to do it. American Journal of Neuroradiology, 41(7), pp.1149–1155.

22. Mafi JN, Edwards ST, Pedersen NP, Davis RB, McCarthy EP, Landon BE. Trends in the ambulatory management of headache: analysis of NAMCS and NHAMCS data 1999-2010. J Gen Intern Med. 2015 May;30(5):548–55. DOI: 10.1007/s11606-014-3107-3. Epub 2015 Jan 8. PMID: 25567755; PMCID: PMC4395605.

23. Kerr, W.T., Tatekawa, H., Lee, J.K., Karimi, A.H., Sreenivasan, S.S., O’Neill, J., Smith, J.M., Hickman, L.B., Savic, I., Nasrullah, N. and Espinoza, R., 2022. Clinical MRI morphological analysis of functional seizures compared to seizure-naïve and psychiatric controls. Epilepsy & Behavior, 134, p.108858.

24. Sheng, L., Ma, H., Shi, Y., Dai, Z., Zhong, J., Chen, F. and Pan, P., 2021. Cortical thickness in migraine: a coordinate-based meta-analysis. Frontiers in neuroscience, 14, p.600423.

25. Henn, A. T., Larsen, B., Frahm, L., Xu, A., Adebimpe, A., Scott, J. C., Linguiti, S., Sharma, V., Basbaum, A. I., Corder, G., Dworkin, R. H., Edwards, R. R., Woolf, C. J., Habel, U., Eickhoff, S. B., Eickhoff, C. R., Wagels, L., & Satterthwaite, T. D. (2022). Structural imaging studies of patients with chronic pain: an anatomical likelihood estimate meta-analysis. Pain, 10.1097/j.pain.0000000000002681. Advance online publication. https://doi.org/10.1097/j.pain.0000000000002681

